# An independent, multi-country head-to-head accuracy comparison of automated chest x-ray algorithms for the triage of pulmonary tuberculosis

**DOI:** 10.1101/2024.06.19.24309061

**Authors:** William Worodria, Robert Castro, Sandra V. Kik, Victoria Dalay, Brigitta Derendinger, Charles Festo, Thanh Quoc Nguyen, Mihaja Raberahona, Swati Sudarsan, Alfred Andama, Balamugesh Thangakunam, Issa Lyimo, Viet Nhung Nguyen, Rivo Rakotoarivelo, Grant Theron, Charles Yu, Claudia M. Denkinger, Simon Grandjean Lapierre, Adithya Cattamanchi, Devasahayam J. Christopher, Devan Jaganath, R2D2 TB Network

**Affiliations:** World Alliance for Lung and Intensive Care in Uganda, Kampala, Uganda; Division of Pulmonary and Critical Care Medicine, University of California, San Francisco, San Francisco, USA; Center for Tuberculosis, University of California, San Francisco, USA; FIND, Geneva, Switzerland; De La Salle Medical and Health Sciences Institute, Dasmarinas Cavite, Philippines; DSI-NRF Centre of Excellence for Biomedical Tuberculosis Research, South African Medical Research Council Centre for Tuberculosis Research, Division of Molecular Biology and Human Genetics, Faculty of Medicine and Health Sciences, Stellenbosch University, South Africa; Ifakara Health Institute, Dar es Salaam, Tanzania; Vietnam National Tuberculosis Programme, National Lung Hospital, Hanoi, Vietnam; VNU University of Medicine and Pharmacy, Hanoi, Vietnam; Department of Infectious Diseases, CHU Joseph Raseta Befelatanana, Antananarivo, Madagascar; Centre d’Infectiologie Charles Mérieux, Université d’Antananarivo, Antananarivo, Madagascar; Department of Pulmonary Medicine, Christian Medical College, Vellore, Tamil Nadu, India; Faculté de Médecine, Université de Fianarantsoa, Fianarantsoa, Madagascar; Department of Infectious Disease and Tropical Medicine, Center for Infectious Diseases, Heidelberg University Hospital, Heidelberg, Germany; German Center for Infection Research, partner site, Heidelberg, Germany; Centre de Recherche du Centre Hospitalier de l’Université de Montréal, Immunopathology Axis, Montréal, Canada; Université de Montréal, Department of Microbiology, Infectious Diseases and Immunology, Montréal, Canada; Division of Pulmonary Diseases and Critical Care Medicine, School of Medicine, University of California Irvine, Orange, USA; Division of Pediatric Infectious Diseases, University of California, San Francisco, San Francisco, USA

**Author notes:** <u>Corresponding author</u> Adithya Cattamanchi, 1001 Health Sciences Road Irvine CA 92697-3950. First authors contributed equally (William Worodria, Robert Castro). Senior authors contributed equally (Devasahayam J. Christopher, Devan Jaganath). See Supplemental Table 1 for additional R2D2 TB Network Contributors.

## Abstract

**Background:** Computer-aided detection (CAD) algorithms for automated chest X-ray (CXR) reading have been endorsed by the World Health Organization for tuberculosis (TB) triage, but independent, multi-country assessment and comparison of current products are needed to guide implementation.

**Methods:** We conducted a head-to-head evaluation of five CAD algorithms for TB triage across seven countries. We included CXRs from adults who presented to outpatient facilities with at least two weeks of cough in India, Madagascar, the Philippines, South Africa, Tanzania, Uganda, and Vietnam. The participants completed a standard evaluation for pulmonary TB, including sputum collection for Xpert MTB/RIF Ultra and culture. Against a microbiological reference standard, we calculated and compared the accuracy overall, by country and key groups for five CAD algorithms: CAD4TB (Delft Imaging), INSIGHT CXR (Lunit), DrAid (Vinbrain), Genki (Deeptek), and qXR (qure.AI). We determined the area under the ROC curve (AUC) and if any CAD product could achieve the minimum target accuracy for a TB triage test (≥90% sensitivity and ≥70% specificity). We then applied country- and population-specific thresholds and recalculated accuracy to assess any improvement in performance.

**Results:** Of 3,927 individuals included, the median age was 41 years (IQR 29-54), 12.9% were people living with HIV (PLWH), 8.2% living with diabetes, and 21.2% had a prior history of TB. The overall AUC ranged from 0.774-0.819, and specificity ranged from 64.8-73.8% at 90% sensitivity. CAD4TB had the highest overall accuracy (73.8% specific, 95% CI 72.2-75.4, at 90% sensitivity), although qXR and INSIGHT CXR also achieved the target 70% specificity. There was heterogeneity in accuracy by country, and females and PLWH had lower sensitivity while males and people with a history of TB had lower specificity. The performance remained stable regardless of diabetes status. When country- and population-specific thresholds were applied, at least one CAD product could achieve or approach the target accuracy for each country and sub-group, except for PLWH and those with a history of TB.

**Conclusions:** Multiple CAD algorithms can achieve or exceed the minimum target accuracy for a TB triage test, with improvement when using setting- or population-specific thresholds. Further efforts are needed to integrate CAD into routine TB case detection programs in high-burden communities.

## INTRODUCTION

Triage tests for pulmonary tuberculosis (TB) are essential to increase access to TB-specific testing and prevent delays in diagnosis and treatment. Globally, an estimated 3.1 of the 10.6 million TB cases are not reported to public health programs each year,^1^ highlighting that missed diagnoses are a major contributor to morbidity, mortality and ongoing transmission. To address this case detection gap, providers and community health workers need the tools to quickly determine who are at higher risk of TB disease to facilitate access to TB-specific testing and treatment initiation.^2^ Ideally, these triage tests should be sensitive, non-invasive and near the point-of-care.^3^ However, there currently is no tool or assay that meets the World Health Organization (WHO) target product profile for a triage test for the general population.

Chest x-ray (CXR) is a sensitive and moderately specific approach to TB triage, but has been limited by the infrastructure and expertise requirements to obtain and interpret the CXR. Computer-aided detection (CAD) algorithms have been developed that utilize deep-learning methods to automatically interpret CXRs with a score output related to the likelihood of TB.^4^ They can further be integrated with digital ultra-portable CXR machines that have limited infrastructure needs.^5^ Several CAD CXR TB products are commercially available,^6^ and overall have shown to be cost-effective with similar performance to human readers.^2,7^ Consequently, the WHO has endorsed CAD algorithms for TB triage in adults.^2^

However, ongoing questions on the performance of CAD algorithms have limited their implementation. The majority of studies have focused on a single CAD platform, preventing head-to-head comparison of each algorithm overall and for key populations including people living with HIV (PLWH) and diabetes. Past studies have also used CXRs obtained with a digital x-ray machine, but current CAD algorithms can also analyze digitized images of CXRs obtained with an analog machine. Multiple analyses have found that the CAD threshold to classify TB may need to be adjusted for different settings and populations, but head-to-head comparisons of CAD products with these thresholds have been limited to one or two countries,^8,9^ retrospective meta-analyses,^10^ or for the screening use-case.^11,12^

An independent, head-to-head comparison of the diagnostic accuracy of CAD algorithms in a large, diverse, multi-country cohort of individuals with presumptive pulmonary TB is needed to address these issues. We thus conducted a prospective diagnostic accuracy study across seven countries in sub-Saharan Africa, South Asia, and Southeast Asia. We independently determined and compared the accuracy of five CAD algorithms to detect pulmonary TB, overall and for key groups, and utilized universal (i.e., single) as well as setting- or population-specific threshold scores.

## METHODS

### Settings and Participants

Participants were enrolled as part of two prospective TB diagnostic accuracy studies, the Rapid Research in Diagnostics Development (R2D2) TB network,^13^ and the Digital Cough Monitoring Project. We included adults 18 years and older with at least two weeks of new or worsening cough from outpatient centers from India, the Philippines, South Africa, Uganda, and Vietnam (R2D2 TB Network), and Madagascar and Tanzania (Digital Cough Monitoring Project) from 2021-2023. We excluded individuals who had completed TB disease or infection treatment in the last 12 months, received antibiotics with anti-mycobacterial activity in the last 2 weeks, or were unable to return for follow-up visits. All participants completed a written informed consent, and the study was approved by the ethical review boards from Christian Medical College (Vellore, India), De La Salle Medical and Health Sciences Institute (Dasmariñas City, Philippines), Stellenbosch University (Cape Town, South Africa), Makerere University College of Health Sciences (Kampala, Uganda), the National Lung Hospital (Hanoi, Vietnam), Comité d’Éthique à la Recheche Biomédicale (Antananarivo, Madagascar), Ifakara Health Institute (Ifakara, Tanzania), the Centre de Recherche du Centre Hospitalier de l’Université de Montréal (Montreal, Canada) and the University of California, San Francisco (San Francisco, USA).

### Procedures

At enrollment, all participants completed a questionnaire on demographics and clinical history, and received a standard TB evaluation by trained personnel. This included an antero-posterior (AP) or postero-anterior (PA) chest X-ray (CXR) and collection of up to three samples of expectorated or induced sputum for *Mycobacterium tuberculosis* complex testing using Xpert MTB/RIF Ultra (Xpert Ultra, Cepheid, Sunnyvale, USA) and mycobacterial culture (liquid or solid) using standard protocols at laboratories by trained staff who were blinded to the CAD results.^14,15^ Individuals enrolled in the R2D2 TB Network returned after three months for follow-up clinical assessment, and repeat CXR and sputum-based mycobacterial testing was repeated if Xpert Ultra testing was negative at baseline.

### CXR Digitization

Digital x-ray machines were available in India, Madagascar, South Africa, Tanzania, and Vietnam. The Philippines site initially used an analog machine retrofitted for digital images, and then transitioned to a digital x-ray machine. An analog machine was used in Uganda until November 2022, and then transitioned to digital x-rays. Research staff were trained at each study site to upload CXRs to a secure cloud-based server. Digital CXRs were in Digital Imaging and Communications in Medicine (DICOM) format, while film-based CXRs were scanned into Joint Photographic Experts Group (JPEG) format. DICOM images had all identifying meta-data removed and JPEG images had all identifying data manually hidden prior to assessment. None of the CXRs had been used previously to train the CAD algorithms.

### CAD Assessment

We independently evaluated five CAD algorithms: CAD4TB version 7 (Delft Imaging, ‘s-Hertogenbosch, the Netherlands), INSIGHT CXR version 3.1.4.1 (Lunit, Seoul, South Korea), qXR version 4 (Qure.AI, Mumbai, India), Genki version 1.1 (DeepTek Medical Imaging Private Limited, Pune, India), and DrAid version 2.0.7-37 (VinBrain, Hanoi, Vietnam). Each CAD software was installed on an online server managed by FIND. CAD analysis was conducted by FIND, according to the developers’ instructions. CAD developers had no access to the images, and no role in the study design, conduct, analysis or interpretation. Each algorithm was then applied to each image, with an output of a TB risk score that ranged from 0-1 (qXR, Genki, DrAid) or 0-100 (CAD4TB, INSIGHT CXR). All CXR images were submitted as DICOM formatted files. Original images in JPEG format were converted into DICOM format using the img2dcm tool from the dcmtk toolkit (v3.6.6). Images that did not fulfill the DICOM features that were required for successful CAD software processing were subsequently modified using the dcmodify tool (v3.6.6) from the dcmtk toolkit before they were processed with the CAD software. The staff performing the assessment were blinded to TB status.

### Reference Standards

Our primary analysis was based on a microbiological reference standard (MRS), defined as TB positive if a participant had a positive baseline Xpert Ultra or culture result, and TB negative if Xpert Ultra negative and at least two negative culture results. Two trace Xpert Ultra results were defined as TB positive. A participant was defined as indeterminate if they had no positive result and less than 2 negative cultures (e.g. due to contamination) and were excluded from the analysis.

### Statistical analyses

We first described the cohort using summary statistics, overall and for each country. Using the CAD TB risk score output, for each algorithm we generated receiver operating characteristic (ROC) curves and calculated the area under each ROC curve (AUC) with 95% confidence intervals (CIs). We determined the threshold that maximizes specificity at 90% sensitivity, to assess if the CAD algorithms could achieve the minimum target accuracy for a TB triage test (≥90% sensitivity and ≥70% specificity). We defined this as the universal threshold as a single cutoff value that could be applied to all countries and subgroups. At the universal threshold, we calculated the sensitivity and specificity with exact binomial 95% CIs of each CAD algorithm, and compared the accuracy of the top-performing algorithm to the other algorithms using McNemar’s test of paired proportions, with significance defined as a p-value < 0.05. We also calculated the accuracy of each algorithm by country and among key subgroups using the universal threshold, including sex, HIV status, diabetes status, and prior history of TB. We generated forest plots to evaluate heterogeneity in country- and group-specific accuracy and assessed if their 95% CIs overlapped with the overall estimate for each CAD algorithm. We then determined if a setting- or population-specific threshold would improve performance by generating ROC curves for each country and subgroup, and calculated the sensitivity and specificity at a threshold that maximized specificity at 90% sensitivity within that group. To enable a head-to-head comparison, we excluded participants who did not have valid results in all CAD platforms, or with indeterminate TB classifications. We presented our findings according to the Standards for Reporting of Diagnostic Accuracy Studies (STARD) criteria.^16^ All analyses were conducted using Stata v. 16.1 (StataCorp, College Station, TX).

## RESULTS

### Participant Characteristics

In total, 4,431 participants were enrolled during the study period and had a baseline CXR analyzed by at least one CAD algorithm (**Figure 1**). Three hundred eight (7%) participants were excluded with indeterminate or missing TB status. Eight (0.2%) were missing a qXR result, 91 (2.1%) had an invalid/error CAD4TB result and 111 (2.5%) had an invalid/error DrAid result. The final number of participants included in the analysis was 3,927, with characteristics described in **Table 1**. The median age was 41 years (interquartile range (IQR) 29-54), 2,133 (54.3%) were male, and 831 (21.2%) had a prior history of TB. The HIV prevalence was 12.9%, and concentrated predominantly in South Africa, Uganda and Tanzania (480/505, 95%). Conversely, 277/3,387 (8.2%) of the cohort had diabetes, based largely in India, the Philippines, and Vietnam (249/277, 89.9%). The microbiological confirmation prevalence was 22.8% (897/3927). About half (56.2%) of those who were Xpert Ultra positive (467/832) had a semi-quantitative level that was medium or high. This proportion was higher in Madagascar (75%) and Uganda (67%), and lower in Tanzania (25%).

**Figure 1.**
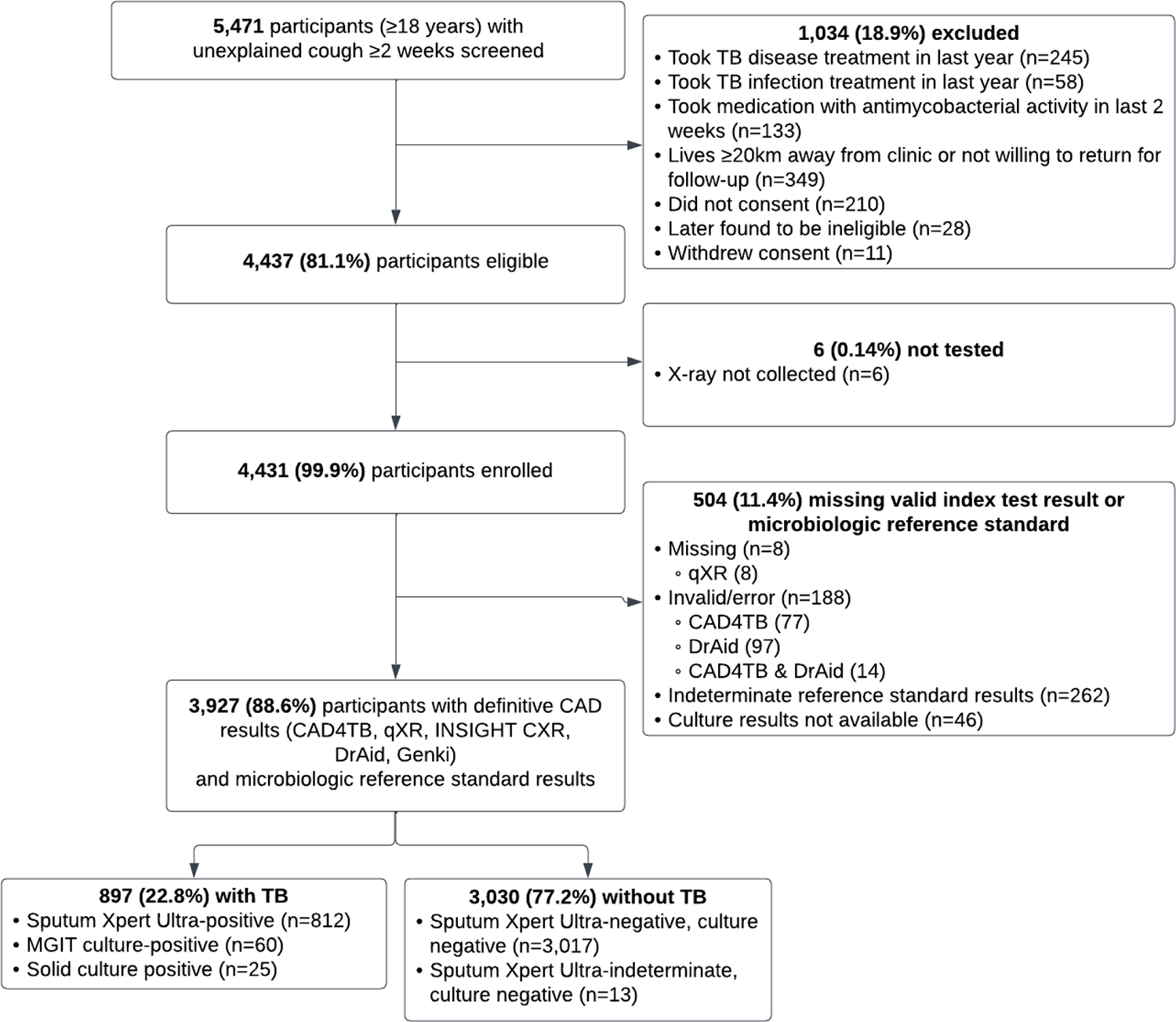
Flowchart of Participants

**Table 1.** Summary of demographic and clinical characteristics, overall and by country.

| **Characteristics**<br>**N (%) unless**<br>**otherwise stated** | **Total** | **Philippines** | **Vietnam** | **South**<br>**Africa** | **Uganda** | **India** | **Tanzania** | **Madagascar** |
| --- | --- | --- | --- | --- | --- | --- | --- | --- |
| **Total in study**<br>**population** | **3,927** | **772 (19.7%)** | **664**<br>**(16.9%)** | **477**<br>**(12.2%)** | **927**<br>**(23.6%)** | **547**<br>**(13.9%)** | **224**<br>**(5.7%)** | **316**<br>**(8.1%)** |
| Age<br>Median (IQR) | 41<br>(29-54) | 41<br>(28-54.5) | 54<br>(40-64) | 38<br>(30-49) | 33<br>(26-42) | 50<br>(36-61) | 42.5<br>(32-52) | 34<br>(25 -50.5) |
| Male | 2,133<br>(54.3%) | 342<br>(44.3%) | 392<br>(59.0%) | 237<br>(49.7%) | 544<br>(58.7%) | 331<br>(60.5%) | 118<br>(52.9%) | 169<br>(53.5%) |
| HIV positive | 505<br>(12.9%) | 4<br>(0.5%) | 4<br>(0.6%) | 176<br>(37.7%) | 232<br>(25.0%) | 14<br>(2.6%) | 72<br>(32.1%) | 3<br>(1.0%) |
| CD4 Count<br>Median (IQR)<sup>1</sup><br>(n=3,387) | 389<br>(194-673) | 356<br>(120-670) | 563<br>(497-597) | 415<br>(214-692) | 350<br>(178-652) | 587<br>(155-737) | - | - |
| Diabetes<sup>1</sup><br>(n=3,387) | 277<br>(8.2%) | 69<br>(8.9%) | 84<br>(12.7%) | 11<br>(2.3%) | 17<br>(1.8%) | 96<br>(17.6%) | - | - |
| Hemoptysis | 564<br>(14.4%) | 53<br>(6.9%) | 135<br>(20.3%) | 32<br>(6.7%) | 159<br>(17.2%) | 76<br>(13.9%) | 38<br>(16.9%) | 71<br>(22.5%) |
| Fever | 1,751<br>(44.6%) | 185<br>(24.0%) | 207<br>(31.2%) | 166<br>(34.8%) | 654<br>(70.6%) | 165<br>(30.2%) | 139<br>(62.1%) | 235<br>(74.4%) |
| Night sweats | 1,563<br>(39.8%) | 162<br>(21.0%) | 153<br>(23.0%) | 268<br>(56.2%) | 573<br>(61.8%) | 84<br>(15.4%) | 117<br>(52.2%) | 206<br>(65.2%) |
| Weight loss | 2,041<br>(52.0%) | 275<br>(35.6%) | 154<br>(23.2%) | 289<br>(60.6%) | 681<br>(73.5%) | 223<br>(40.8%) | 133<br>(59.4%) | 286<br>(90.5%) |
| Poor appetite<sup>1</sup><br>(n=3,387) | 1,240<br>(36.6%) | 236<br>(30.6%) | 121<br>(18.2%) | 191<br>(40.0%) | 488<br>(52.6%) | 204<br>(37.3%) | - | - |
| Lymphadenopathy*<br>(n=3,387) | 150<br>(4.4%) | 20<br>(2.6%) | 7<br>(1.0%) | 24<br>(5.0%) | 95<br>(10.3%) | 4<br>(0.7%) | - | - |
| History of TB | 831<br>(21.2%) | 203<br>(26.3%) | 158<br>(23.8%) | 154<br>(32.3%) | 127<br>(13.7%) | 80<br>(14.6%) | 63<br>(28.1%) | 46<br>(14.6%) |
| History of contact*<br>(n=3,387) | 818<br>(24.2%) | 366<br>(47.4%) | 45<br>(6.8%) | 103<br>(21.6%) | 254<br>(27.4%) | 50<br>(9.1%) | - | - |
| History of smoking<br>(last 7 days) | 798<br>(20.3%) | 245<br>(31.7%) | 99<br>(14.9%) | 223<br>(46.8%) | 107<br>(11.5%) | 28<br>(5.1%) | 33<br>(14.7%) | 63<br>(19.9%) |
| Microbiologically-<br>confirmed TB | 897<br>(22.8%) | 82<br>(10.6%) | 201<br>(30.3%) | 90<br>(18.9%) | 308<br>(33.2%) | 50<br>(9.1%) | 35<br>(15.6%) | 131<br>(41.5%) |
| Xpert Ultra positive | 832<br>(21.2%) | 70<br>(9.1%) | 187<br>(28.2%) | 84<br>(17.6%) | 297<br>(32.0%) | 49<br>(9.0%) | 24<br>(10.7%) | 121<br>(38.3%) |
| Trace | 32<br>(3.9%) | 2<br>(2.9%) | 8<br>(4.3%) | 7<br>(8.3%) | 7<br>(2.4%) | 4<br>(8.2%) | 1<br>(4.2%) | 3<br>(2.5%) |
| Very Low | 101<br>(12.1%) | 16<br>(22.9%) | 28<br>(15.0%) | 13<br>(15.5%) | 23<br>(7.7%) | 11<br>(22.5%) | 5<br>(20.8%) | 5<br>(4.1%) |
| Low | 232<br>(27.9%) | 24<br>(34.3%) | 68<br>(36.4%) | 20<br>(23.8%) | 68<br>(22.9%) | 18<br>(36.7%) | 12<br>(50.0%) | 22<br>(18.2%) |
| Medium | 197<br>(23.7%) | 15<br>(21.4%) | 42<br>(22.5%) | 24<br>(28.6%) | 84<br>(28.3%) | 10<br>(20.4%) | 2<br>(8.3%) | 20<br>(16.5%) |
| High | 270<br>(32.5%) | 13<br>(18.6%) | 41<br>(21.9%) | 20<br>(23.8%) | 115<br>(38.7%) | 6<br>(12.2%) | 4<br>(16.7%) | 71<br>(58.7%) |

IQR: interquartile range; TB: tuberculosis
1. Data unavailable from Tanzania and Madagascar, and denominator indicated

### Head-to-head comparison of CAD algorithm accuracy

The ROC curves for each algorithm are shown in **Figure 2**. The AUCs were similar across CAD algorithms, ranging from 0.774-0.819. At 90% sensitivity, CAD4TB had the highest specificity at 73.8% (95% CI 72.2-75.4), although qXR and INSIGHT CXR also achieved the minimum target of 70% specificity (**Table 2**) with similar AUCs across the three products (0.800-0.819). DrAid and Genki were less specific, at 67.9% and 64.8%, respectively. In pairwise comparison, CAD4TB was significantly more specific than the other algorithms (p<0.001), although the absolute difference ranged from 3.5-9% (**Table 2**).

**Figure 2.**
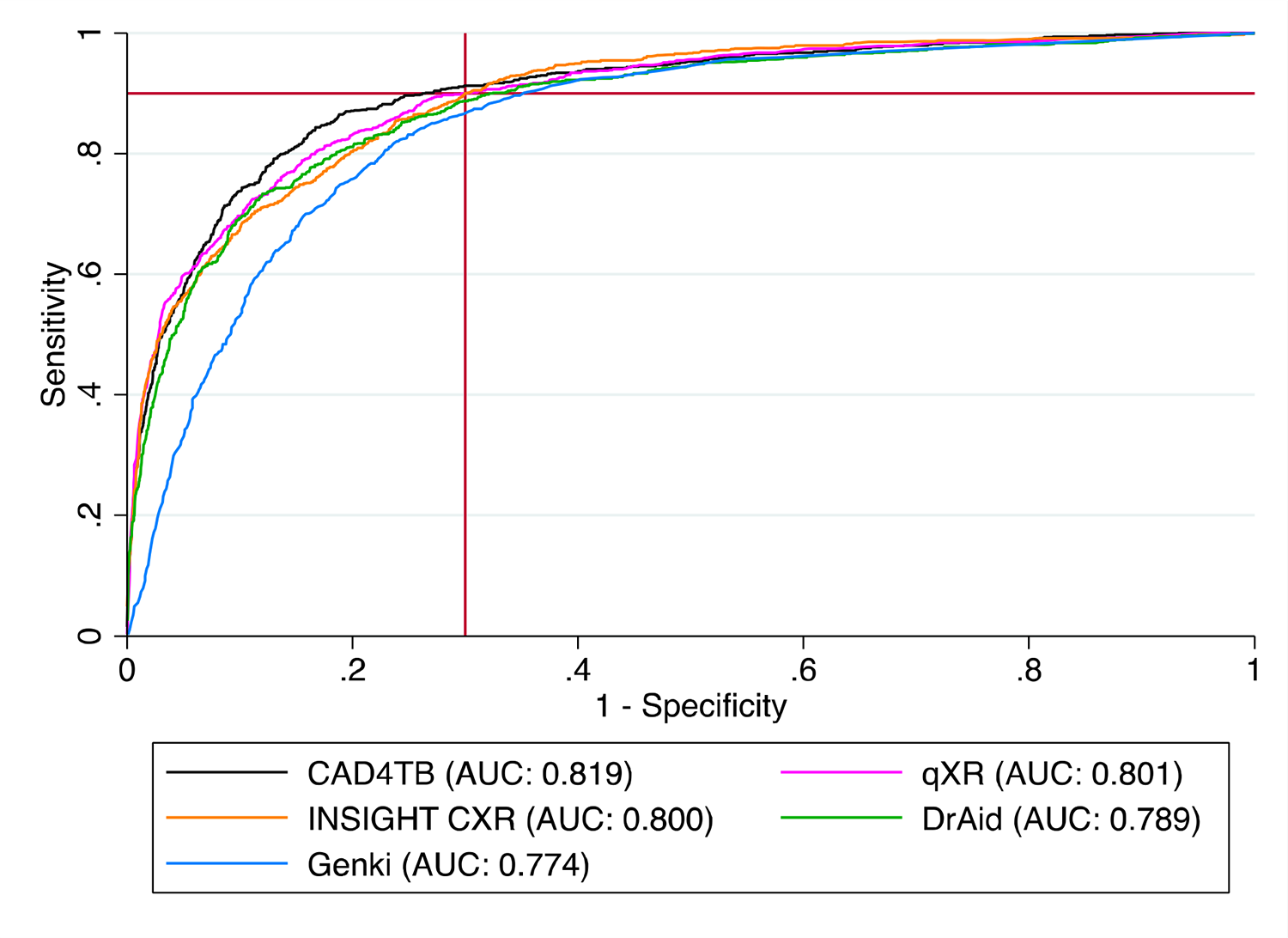
Receiver operating characteristic curve of each CAD Algorithm. Each ROC curve represents a CAD algorithm as indicted in the legend, with reported area under the curve (AUC). The red horizontal and vertical lines indicate minimum target sensitivity and specificity for a TB triage test at 90% and 70%, respectively.

**Table 2.** Head-to-head accuracy of each CAD algorithm.

| **CAD Algorithm** | **AUC (95% CI)** | **Threshold of positivity<sup>1</sup>** | **Sensitivity % (95% CI)<sup>2</sup>** | **Specificity % (95% CI)** | **Difference in Specificity vs. CAD4TB % (95% CI)** | **p-value** |
| --- | --- | --- | --- | --- | --- | --- |
| **CAD4TB** | 0.819<br>(0.806-0.831) | ≥36.31 | 90%<br>(87.8-91.9) | 73.8%<br>(72.2-75.4) | - | - |
| **qXR** | 0.801<br>(0.789-0.814) | ≥0.289 | 90%<br>(87.8-91.9) | 70.3%<br>(68.7-72.0) | 3.5%<br>(2.2%, 4.8%) | < 0.001 |
| **INSIGHT CXR** | 0.800<br>(0.787-0.813) | ≥8.25 | 90%<br>(87.8-91.9) | 70.0%<br>(68.4-71.7) | 3.8%<br>(2.3%, 5.2%) | < 0.001 |
| **DrAid** | 0.789<br>(0.776-0.802) | ≥0.2149 | 90%<br>(87.8-91.9) | 67.9%<br>(66.2-69.5) | 5.9%<br>(4.3%, 7.5%) | < 0.001 |
| **Genki** | 0.774<br>(0.762-0.787) | ≥0.06667 | 90.1%<br>(87.9-92.0) | 64.8%<br>(63.1-66.5) | 9.0%<br>(7.4%, 10.5%) | < 0.001 |

AUC: Area under the receiver operating characteristic (ROC) curve; CAD: Computer-Aided Detection
1. TB risk scores ranged from 0-100 for CAD4TB and INSIGHT CXR, and 0-1 for qXR, DrAid and Genki 2. Threshold based on a target sensitivity of 90%, and calculated on the total dataset (defined as “universal threshold”)

### The accuracy of CAD algorithms by country and subgroup – Universal threshold

When stratified by country, we found heterogeneity in accuracy as shown in **Figure 3A** for the highest performing algorithm (CAD4TB) and in **Supplemental Figures 1A-4A** for the other algorithms. For CAD4TB, using the universal calculated threshold score of 36.31, sensitivity ranged from 80% to 95.5%, although the 95% CIs of each country overlapped or exceeded the overall estimate of 90%. Specificity ranged from 67% to 83.6%, and was reduced in Vietnam and Madagascar. South Africa was the only country achieving the minimum target accuracy for a TB triage test with CAD4TB (Sensitivity 93.3% (95% CI 86.1-97.5) and specificity 71.6% (95% CI 66.8-76)) when using the universal threshold. Across the other algorithms, performance remained similar to the overall estimates of each CAD product in the Philippines, India and Tanzania. Specificity at the universal threshold was generally lower than the overall estimate in Vietnam for qXR and DrAid, but was improved with INSIGHT CXR and Genki. In Uganda, sensitivity was lower by qXR and INSIGHT CXR, and specificity was lower with DrAid. In South Africa, specificity was marginally reduced with INSIGHT CXR and Genki. In Madagascar, specificity was generally lower than the overall estimate for each CAD product except for DrAid. In India, specificity was improved with DrAid.

**Figure 3.**
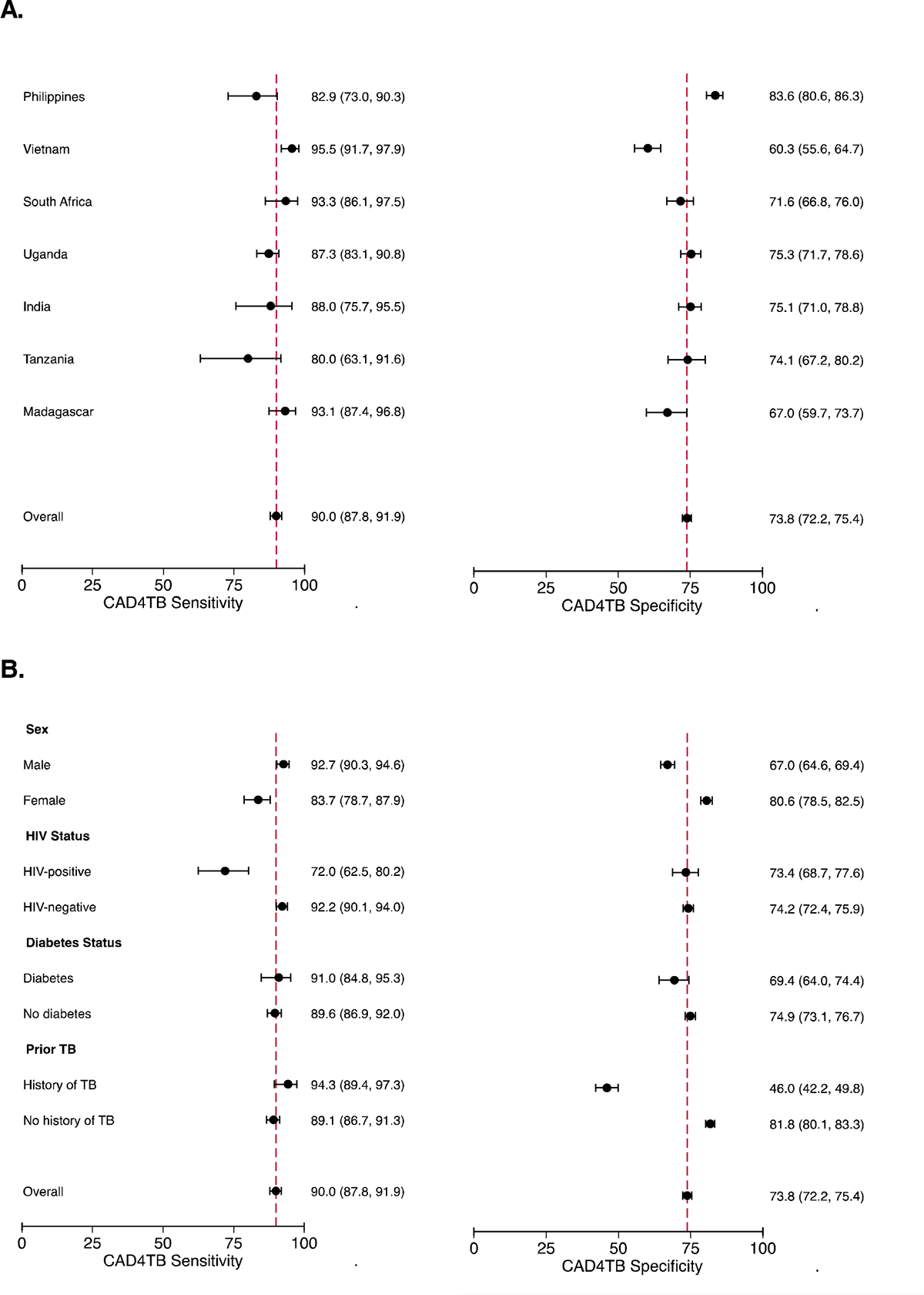
Forest plot of the sensitivity and specificity of CAD4TB by country and subgroup using a universal threshold. (A) The sensitivity and specificity by country, with 95% CIs; (B) The sensitivity and specificity by subgroup, with 95% CIs. The overall accuracy of the CAD algorithm is listed at the bottom with a vertical dashed red line, in order to compare the overall estimate to the country and subgroup estimates.

**Figure 4.**
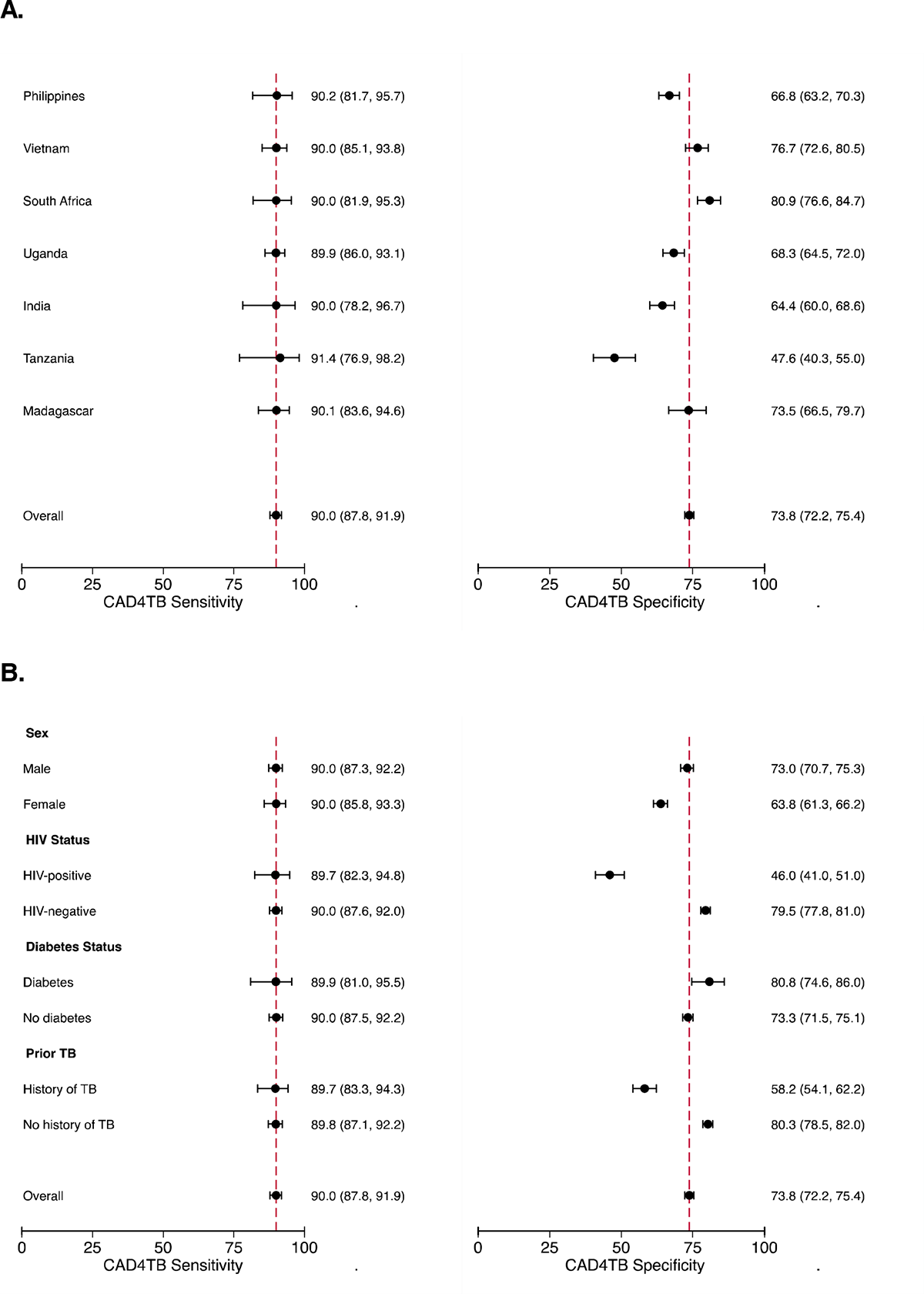
Forest plot of the sensitivity and specificity of CAD4TB by country and subgroup using country- and population-specific thresholds. (A) The sensitivity and specificity by country, with 95% CIs; and (B) The sensitivity and specificity by subgroup, with 95% CIs. Of note, the threshold selected is based on a 90% sensitivity. The overall accuracy of the CAD algorithm is listed at the bottom with a vertical dashed red line, in order to compare the overall estimate to the country and subgroup estimates.

We also found heterogeneity when the accuracy was assessed in key subgroups using the universal threshold (**Figure 3B** for CAD4TB, and **Supplemental Figures 1B-4B** for other algorithms). For CAD4TB, sensitivity was lower in females and people living with HIV (PLWH), while specificity was lower in males and those with a history of TB compared to the overall estimates. Sensitivity in people living with diabetes (PLWD) was similar to those without diabetes; specificity was slightly reduced to 69.4% (95% CI 64-74.4) in PLWD although still close to the minimum target accuracy. Trends were similar across algorithms, with generally lower sensitivity in females and PLWH, and lower specificity in males and those with a history of TB. There was no heterogeneity by diabetes status.

### Application of Population-specific Thresholds

As shown in **Figure 4** for CAD4TB and **Supplemental Table 2** for other algorithms, we applied country- and population-specific thresholds and determined the specificity at 90% sensitivity. Among countries that had a CAD4TB sensitivity of less than 90% (Philippines, Uganda, India, and Tanzania), increasing sensitivity with a country-specific threshold resulted in a lower specificity. For the Philippines, Uganda and India, the specificity remained within 10% of the minimum target accuracy of 70% and ranged from 64.4-68.3%. Tanzania had the lowest sensitivity initially (80%) with CAD4TB, and so increasing its sensitivity to 90% lowered the specificity to 47.6% (95% CI 40.3-55). For Vietnam, South Africa, and Madagascar that had greater than 90% sensitivity when using the universal threshold, lowering the sensitivity allowed all three to exceed the minimum target specificity (range 76.7-80.9%). For most countries, at least one CAD product achieved the minimum target accuracy for a TB triage test. The specificity in Uganda was close to the target accuracy, with specificity ranging from 68.3-68.8% for CAD4TB, qXR and INSIGHT CXR. In Tanzania, qXR had the highest specificity of 64% (95% CI 56.7-70.9) at 90% sensitivity. INSIGHT CXR achieved the minimum target accuracy for a TB triage test for the greatest number of countries (5/7).

When group-specific thresholds were applied, the minimum target accuracy could be achieved or exceeded with CAD4TB for males, people without HIV, people with and without diabetes and people without history of TB. Increasing the sensitivity to 90% reduced the specificity of CAD4TB among females to 63.8% (95% CI 61.3-66.2) and PLHW to 46% (95% CI 41-51). A male-specific threshold improved the specificity to 73% (95% CI 70.7-75.3); however, a subgroup specific threshold for people with a history of TB was unable to substantially improve specificity which remained low (58.2%, 95% CI 54.1-62.2). Similar trends were seen in other algorithms. The highest specificity for females was with INSIGHT CXR, where females achieved close to the target accuracy at 68.8% specificity (95% CI 66.4-71.1), while PLWH reached 53% specificity (95% CI 48-58) at 90% sensitivity with qXR. CAD4TB achieved the highest specificity for people with a history of TB at 58.2%. CAD4TB was able to achieve or exceed the minimum target accuracy for a TB triage test for the greatest number of groups assessed (5/8).

## DISCUSSION

Automated CXR reading with CAD algorithms have provided an innovative tool to support the triage of individuals being evaluated for pulmonary TB. With several commercial products available, clinical and public health programs need to decide which algorithm(s) to implement. We performed a large independent head-to-head assessment of CAD products across seven countries, and found that overall accuracy was similar and CAD4TB, qXR and INSIGHT CXR achieved the minimum WHO target accuracy for a TB triage test. There was heterogeneity in accuracy by country and among key subgroups that was overall similar across CAD algorithms; however, application of country- and population-specific thresholds achieved or approached the minimum target accuracy for at least one CAD product, though gaps remained among PLWH and those with a history of TB. These finding demonstrate that there are multiple CAD options that are valuable for TB triage, with good performance across countries and subgroups that can be further fine-tuned according to local demographics.

The overall accuracy was comparable across CAD products, with CAD4TB having the highest specificity followed by qXR and INSIGHT CXR. This is similar to an individual patient data (IPD) meta-analysis of studies from four countries that found similar performance across CAD4TB, qXR and INSIGHT CXR.^10^ Specificity was lower in that study (ranging 54-61% specificity at 90% sensitivity),^10^ although older CAD versions were used in that study and have been shown to not perform as well as current algorithms.^9^ It is encouraging that the current algorithms can achieve the minimum target accuracy for a TB triage test. One study compared Genki to other CAD algorithms and noted similar specificity to CAD4TB and qXR, while we found it to be overall less specific.^11^ However, that study assessed CAD in a screening cohort and was conducted in Vietnam where we also found Genki had higher specificity, highlighting the importance to conduct a multi-country evaluation to assess performance. To our knowledge this is the first published work to assess and compare DrAid, and although lower accuracy than the above three algorithms, overall it performed well with 68% specificity at 90% sensitivity. While CAD4TB was the highest performing algorithm, it should be noted that other studies have found it to be similar to INSIGHT CXR and qXR,^8,11,17^ and CAD4TB had more invalid or error results. Our findings overall demonstrate that there are several CAD algorithms that can now achieve the minimum target accuracy for a TB triage test when compared across multiple countries and regions.

When assessed by country and population, CAD performance was heterogenous. This has been well-described by previous studies that have compared CAD4TB, qXR and INSIGHT CXR and have found that accuracy varied by country, and was lower for females, PLWH, and history of TB.^8,10–12,17,18^ Few studies have assessed CAD for PLWD; screening studies in Indonesia and Pakistan found that specificity was low at 17-42% at about 90% sensitivity for CAD4TB.^19,20^ A separate study in Pakistan found that INSIGHT CXR had similar performance among those with and without diabetes (87% sensitivity and 60-64% specificity).^21^ We found that the accuracy was stable among those with and without diabetes, and is encouraging that there are several CAD products that perform well for this at-risk population, especially in TB endemic regions with a higher diabetes prevalence such as South and Southeast Asia. Variation in CAD product performance by setting and subgroup likely reflects the methods and population used to train the models.^8,12^ Differences between country cohorts may also explain differences in accuracy; for example, sensitivity was reduced in Tanzania where 75% had lower bacterial burden by Xpert semi-quantitative level. However, in South Africa which had a large proportion of people living with HIV and with a prior history of TB, performance was overall stable across algorithms.

To address the heterogeneity, we applied country- or population-specific thresholds, and found that at least one CAD product could achieve or was close to the minimum target accuracy for a TB triage test for each country and most groups. This was an improvement in comparison to the IPD meta-analysis that was unable to substantially increase performance with country-specific thresholds.^10^ The exceptions were PLWH and those with a history of TB, likely due to the low sensitivity to detect lung abnormalities in PLWH who have paucibacillary disease, and low specificity among those with a history of TB given persistent abnormalities on imaging. It is important to note that similar variation has been seen in human readers of CXRs for TB,^10,22^ and so there is still potential value in settings where providers do not have access to expert CXR reads and for improved reliability.

Our findings can help support programmatic decision-making in the implementation of CAD algorithms. In our multi-country analysis, there are currently several CAD algorithms available that could be utilized based on accuracy and consideration of the local demographics. Facilities and TB programs can consider then other factors including cost and infrastructure needs for each product. Moreover, each product may have other features that may be desirable to the program; for example, the CAD4TB version we evaluated provided an output of TB score and classification, while the other algorithms also indicated other abnormalities.^6^ Regardless of the CAD algorithm, our findings support that current CAD products may need threshold adjustment prior to implementation. The WHO has developed a toolkit to guide local calibration,^23^ and may be further supported by some of the CAD products. The thresholds we identified may be useful as a starting point, although updated versions of CAD algorithms may require re-assessment. Moreover, the chosen threshold should also be guided by the main goals of the program, balancing reduction in confirmatory testing with risk of missed cases, and considerations of cost-effectiveness.^7,8,24^

Our study independently assessed the accuracy of multiple CAD products in the greatest number of countries to date, overall and among key risk groups. We also included two algorithms (DrAid and Genki) that have not been compared in the triage use-case previously. CXRs were obtained from well-characterized cohorts, with a microbiological reference standard that included culture to increase yield beyond Xpert alone. Previous studies have assessed digital CXRs alone, while our study included a mix of digital and analog images. There were some limitations. We did not compare CAD products to a human interpretation, which requires a panel of expert readers and standardized annotation given high inter-reader variability. This was outside the scope of our study, and has been well-assessed previously.^8,10,11^ All participants had cough, and we would have benefited from including individuals who did not have cough and met other screening criteria for TB testing. Some data was not available in Tanzania and Madagascar, including diabetes status, which may have biased assessment of heterogeneity, although there was still East African representation from Uganda. We were not powered to assess threshold identification by both country and subgroup, though as above the threshold should be further guided by the overall demographics and goals of the program. CAD algorithms continue to be developed or optimized with new versions, and these will require future independent validation.^25^

## CONCLUSIONS

Across seven countries in high TB-burden settings, we found that there are several CAD algorithms that achieved the WHO target accuracy for a TB triage test. The CAD products can be further tuned to achieve goal accuracy depending on the key demographics of interest. Further work is needed to improve performance in PLWH and those with a history of TB, including in combination with other triage tests. Thus, CAD for automated CXR reading has large potential to expand TB diagnosis and treatment globally, with greater focus now needed on the implementation factors to increase access to high-burden communities.

## Data Availability

All data produced in the present study are available upon reasonable request to the authors

## ACKNOWLEDGEMENTS

We acknowledge the patients who participated, and all the R2D2 TB Network and Digital Cough Monitoring study personnel.

## CONFLICTS OF INTEREST

The authors declare no conflicts of interest.

The installation and use of the different CAD software evaluated in this manuscript was provided free of charge by all CAD vendors to FIND. CAD vendors did not have any role in the study design, data collection, analysis, the decision to publish or the preparation of the manuscript.

