## Supplemental for "An independent, multi-country head-to-head accuracy comparison of automated chest x-ray algorithms for the triage of pulmonary tuberculosis"

### SUPPLMENTAL TABLES AND FIGURES

#### Supplemental Table 1. Additional contributors from the R2D2 TB Network

| First name | Surname | Affiliation |
|---|---|---|
| **India** | | |
| Shanmugasundaram | Elango | Christian Medical College, Vellore, India |
| Jerusha | Emmanuel | Christian Medical College, Vellore, India |
| Vinita | Ernest | Christian Medical College, Vellore, India |
| Priyadarshini | Gajendran | Christian Medical College, Vellore, India |
| Flavita | John | Christian Medical College, Vellore, India |
| Bharath | Karthikeyan | Christian Medical College, Vellore, India |
| Divya | Mangal | Christian Medical College, Vellore, India |
| Swetha | Sankar | Christian Medical College, Vellore, India |
| Rajasekar | Sekar | Christian Medical College, Vellore, India |
| Reena | Sekar | Christian Medical College, Vellore, India |
| Deepa | Shankar | Christian Medical College, Vellore, India |
| Mary | Shibiya | Christian Medical College, Vellore, India |
| Sai | Vijayasree | Christian Medical College, Vellore, India |
| **Philippines** | | |
| Jared | Almonte | De La Salle Medical and Health Sciences Institute, Cavite |
| Kevin Joshua | Alonzo | National TB Reference Laboratory, Research Institute for Tropical Medicine, Department of Health |
| Mary Faith | Angcaya | De La Salle Medical and Health Sciences Institute, Cavite |
| Joseph Edwin L. | Bascuña | National TB Reference Laboratory, Research Institute for Tropical Medicine, Department of Health |
| Ramon P. | Basilio | National TB Reference Laboratory, Research Institute for Tropical Medicine, Department of Health |
| Asella Ruvijean | Cariaga | De La Salle Medical and Health Sciences Institute, Cavite, Philippines |
| Gabriella | Castillon | De La Salle Medical and Health Sciences Institute, Cavite |
| Karlo | Dayawon | De La Salle Medical and Health Sciences Institute, Cavite |
| Raul | Destura | National Institutes of Health, University of the Philippines Manila |
| Jezreel | Esguerra | De La Salle Medical and Health Sciences Institute, Cavite |
| Eleonor | Garcia | De La Salle Medical and Health Sciences Institute, Cavite |
| Darecil | Gelina | De La Salle Medical and Health Sciences Institute, Cavite |
| Joseph Aldwin | Goleña | De La Salle Medical and Health Sciences Institute, Cavite |

| First name | Surname | Affiliation |
|---|---|---|
| Maria Marissa | Golla | De La Salle Medical and Health Sciences Institute, Cavite |
| Emmanuelle | Gutierrez | De La Salle Medical and Health Sciences Institute, Cavite |
| Gidalthi Jonathan | Ilagan | De La Salle Medical and Health Sciences Institute, Cavite |
| Dodge R. | Lim | National TB Reference Laboratory, Research Institute for Tropical Medicine, Department of Health |
| Jaiem | Maranan | De La Salle Medical and Health Sciences Institute, Cavite |
| Danaida | Marcelo | De La Salle Medical and Health Sciences Institute, Cavite |
| Leonedy | Masangcay | De La Salle Medical and Health Sciences Institute, Cavite |
| Jenkin | Mendoza | National TB Reference Laboratory, Research Institute for Tropical Medicine, Department of Health |
| Angelita | Pabruada | De La Salle Medical and Health Sciences Institute, Cavite |
| Laarean | Perlas | De La Salle Medical and Health Sciences Institute, Cavite |
| Annalyn | Reyes | De La Salle Medical and Health Sciences Institute, Cavite |
| Roeus Vincent Arjay G. | Reyes | National TB Reference Laboratory, Research Institute for Tropical Medicine, Department of Health |
| Lorenzo | Reyes | National TB Reference Laboratory, Research Institute for Tropical Medicine, Department of Health |
| Maria Guileane | Sanchez-Pogosa | National TB Reference Laboratory, Research Institute for Tropical Medicine, Department of Health |
| Maricef | Tonquin | De La Salle Medical and Health Sciences Institute, Cavite |
| **South Africa** | | |
| Shima | Abdulgadar | Stellenbosch University, Cape Town, South Africa |
| Cammy | Botha | Stellenbosch University, Cape Town, South Africa |
| Welile | Dube-Nwamba | Stellenbosch University, Cape Town, South Africa |
| Jane | Fortuin | Stellenbosch University, Cape Town, South Africa |
| Siphosethu | Gonya | Stellenbosch University, Cape Town, South Africa |
| Chumani | Hatile | Stellenbosch University, Cape Town, South Africa |

| First name | Surname | Affiliation |
|---|---|---|
| Megan | Hendrikse | Stellenbosch University, Cape Town, South Africa |
| Charlotte | Lawn | Stellenbosch University, Cape Town, South Africa |
| Disha | Mathoorah | Stellenbosch University, Cape Town, South Africa |
| Desiree Lem | Mbu | Stellenbosch University, Cape Town, South Africa |
| Zintle | Ntetha | Stellenbosch University, Cape Town, South Africa |
| Anna | Okunola | Stellenbosch University, Cape Town, South Africa |
| Zaida | Palmer | Stellenbosch University, Cape Town, South Africa |
| Fikiswa | Seti | Stellenbosch University, Cape Town, South Africa |
| Charmaine | Van Der Walt | Stellenbosch University, Cape Town, South Africa |
| Lusanda | Yekani | Stellenbosch University, Cape Town, South Africa |
| **Uganda** | | |
| Lucy | Asege | Walimu, Kampala, Uganda |
| Alice | Bukirwa | Walimu, Kampala, Uganda |
| David | Katumba | Walimu, Kampala, Uganda |
| Esther | Kisakye | Walimu, Kampala, Uganda |
| Wilson | Mangeni | Walimu, Kampala, Uganda |
| Job | Mukwatamundu | Walimu, Kampala, Uganda |
| Sandra | Mwebe | Walimu, Kampala, Uganda |
| Annet | Nakaweesa | Walimu, Kampala, Uganda |
| Martha | Nakaye | Walimu, Kampala, Uganda |
| Talemwa | Nalugwa | Walimu, Kampala, Uganda |
| Irene | Nassuna | Walimu, Kampala, Uganda |
| Irene | Nekesa | Walimu, Kampala, Uganda |
| Justine | Nyawere | Walimu, Kampala, Uganda |
| John Baptist | Ssonko | Walimu, Kampala, Uganda |
| **Vietnam** | | |
| Hai | Dang | Vietnam National Tuberculosis Program-University of California San Francisco Research Collaboration Unit; Center for Promotion of Advancement of Society, Hanoi, Vietnam |
| Luong | Dinh | Vietnam National Lung Hospital |
| Hang | Do | Hanoi Lung Hospital, Hanoi, Vietnam |
| Tam | Do | Hanoi Lung Hospital, Hanoi, Vietnam |
| Thuong | Do | Vietnam National Lung Hospital |
| Dung | Dao | Hanoi Lung Hospital, Hanoi, Vietnam |
| Ha | Doan | National TB reference Lab/ Vietnam National Lung Hospital, Hanoi, Vietnam |
| Thien | Doan | Hanoi Lung Hospital, Hanoi, Vietnam |

| First name | Surname | Affiliation |
|---|---|---|
| Huy | Ha | Vietnam National Tuberculosis Program-University of California San Francisco Research Collaboration Unit, Center for Promotion of Advancement of Society, Hanoi, Vietnam |
| Oanh | Lai | Hanoi Lung Hospital, Hanoi, Vietnam |
| Hien | Le | Vietnam National Tuberculosis Program-University of California San Francisco Research Collaboration Unit; Center for Promotion of Advancement of Society, Hanoi, Vietnam |
| Nguyet | Le | National TB reference Lab/ Vietnam National Lung Hospital, Hanoi, Vietnam |
| Anh | Nguyen | Hanoi Lung Hospital, Hanoi, Vietnam |
| Dong | Nguyen | Hanoi Lung Hospital, Hanoi, Vietnam |
| Hanh | Nguyen | Vietnam National Tuberculosis Program-University of California San Francisco Research Collaboration Unit; Center for Promotion of Advancement of Society, Hanoi, Vietnam |
| Hoa | Nguyen | Vietnam National Lung Hospital |
| Hoang | Nguyen | Hanoi Lung Hospital, Hanoi, Vietnam |
| Yen | Nguyen | Hanoi Lung Hospital, Hanoi, Vietnam |
| Ha | Phan | Vietnam National Tuberculosis Program-University of California San Francisco Research Collaboration Unit, Center for Promotion of Advancement of Society, Hanoi, Vietnam |
| Nam | Pham | Vietnam National Tuberculosis Program-University of California San Francisco Research Collaboration Unit, Hanoi Lung Hospital, Hanoi, Vietnam |
| Thuong | Pham | Hanoi Lung Hospital, Hanoi, Vietnam |
| Trang | Trinh | Vietnam National Tuberculosis Program-University of California San Francisco Research Collaboration Unit, Center for Promotion of Advancement of Society, Hanoi, Vietnam |
| Phuong | Vu | Hanoi Lung Hospital, Hanoi, Vietnam |
| Trung | Vu | National TB reference Lab/ Vietnam National Lung Hospital, Hanoi, Vietnam |
| **USA** | | |
| Catherine | Cook | University of California San Francisco, San Francisco, CA, USA |
| Rebecca | Crowder | University of California San Francisco, San Francisco, CA, USA |
| Sophie | Huddart | University of California San Francisco, San Francisco, CA, USA |
| Midori | Kato-Maeda | University of California San Francisco, San Francisco, CA, USA |
| Tessa | Mochizuki | University of California San Francisco, San Francisco, CA, USA |
| Ruvandhi | Nathavitharana | Beth Israel Deaconess Medical Center, Harvard Medical School, Boston, MA, USA |

| First name | Surname | Affiliation |
|---|---|---|
| Payam | Nahid | University of California San Francisco, San Francisco, CA, USA |
| Kevin | Nolan | University of California San Francisco, San Francisco, CA, USA |
| Patrick | Phillips | University of California San Francisco, San Francisco, CA, USA |
| Kinari | Shah | University of California San Francisco, San Francisco, CA, USA |
| Christina | Yoon | University of California San Francisco, San Francisco, CA, USA |
| **Germany** | | |
| Maria del Mar | Castro Noriega | Heidelberg University Hospital, Heidelberg, Germany |
| Claudia | Denkinger | Heidelberg University Hospital, Heidelberg, Germany |
| Ankur | Gupta-Wright | Heidelberg University Hospital, Heidelberg, Germany<br>Department of Infectious Diseases, Imperial College London, UK |
| Theresa | Pfurtscheller | Heidelberg University Hospital, Heidelberg, Germany |
| Seda | Yerlikaya | Heidelberg University Hospital, Heidelberg, Germany |
| **Switzerland** | | |
| Matthew | Arentz | FIND, Geneva, Switzerland |
| Nathalie | Frey | FIND, Geneva, Switzerland |
| Sandra V. | Kik | FIND, Geneva, Switzerland |
| Sam | Linsen | FIND, Geneva, Switzerland |
| Morten | Ruhwald | FIND, Geneva, Switzerland |
| **Tanzania** | | |
| Rose | Chagama | Temeke Regional Referral Hospital, Dar es Salaam, Tanzania |
| Lydia | J. Balehetse | Ifakara Health Institute, Dar es Salaam, Tanzania |
| Gasper | Kimaro | Temeke Regional Referral Hospital, Dar es Salaam, Tanzania |
| Omar | Lweno | Ifakara Health Institute, Dar es Salaam, Tanzania |
| Antelius | M. Ngatulile | Ifakara Health Institute, Dar es Salaam, Tanzania |
| Husna | Msangi | Temeke Regional Referral Hospital, Dar es Salaam, Tanzania |
| **Madagascar** | | |
| Niaina | Rakotosamimanana | Institut Pasteur de Madagascar, Antananarivo, Madagascar |
| **Canada** | | |
| Dominique Hélène | Doré | Centre de Recherche du Centre Hospitalier de l'Université de Montréal, Immunopathology Axis, Montréal, Canada |

**Supplemental Table 2. Specificity of each CAD algorithm at 90% sensitivity, using country- and population-specific thresholds**

| | CAD4TB | | qXR | | INSIGHT CXR | | DrAid | | Genki | |
|---|---|---|---|---|---|---|---|---|---|---|
| | **Specificity at 90% sensitivity, % (95% CI)** | **Threshold of positivity** | **Specificity at 90% sensitivity, % (95% CI)** | **Threshold of positivity** | **Specificity at 90% sensitivity, % (95% CI)** | **Threshold of positivity** | **Specificity at 90% sensitivity, % (95% CI)** | **Threshold of positivity** | **Specificity at 90% sensitivity, % (95% CI)** | **Threshold of positivity** |
| Universal Threshold | 73.8% (72.2, 75.4) | ≥36.31 | 70.3% (68.7, 72.0) | ≥0.289 | 70.0% (68.4, 71.7) | ≥8.25 | 67.9% (66.2, 69.5) | ≥0.2149 | 64.8% (63.1, 66.5) | ≥0.06667 |
| **Country** | | | | | | | | | | |
| Philippines | 66.8% (63.2%, 70.3%) | ≥14.65 | 78.1% (74.8%, 81.1%) | ≥0.566 | 74.6% (71.2%, 77.8%) | ≥13.24 | 79.4% (76.2%, 82.4%) | ≥0.2846 | 66.8% (63.2%, 70.3%) | ≥0.05098 |
| Vietnam | 76.7% (72.6%, 80.5%) | ≥54.92 | 77.3% (73.2%, 81.1%) | ≥0.684 | 81.2% (77.3%, 84.7%) | ≥22.37 | 74.7% (70.5%, 78.6%) | ≥0.5198 | 67.4% (62.9%, 71.6%) | ≥0.07843 |
| South Africa | 80.9% (76.6%, 84.7%) | ≥50.65 | 76.0% (71.4%, 80.1%) | ≥0.442 | 71.8% (67.1%, 76.3%) | ≥17.64 | 58.9% (53.8%, 63.9%) | ≥0.1319 | 70.5% (65.7%, 75.0%) | ≥0.27451 |
| Uganda | 68.3% (64.5%, 72.0%) | ≥30.16 | 68.7% (64.8%, 72.3%) | ≥0.131 | 68.8% (65.0%, 72.5%) | ≥3.46 | 54.0% (49.9%, 57.9%) | ≥0.1696 | 64.9% (61.0%, 68.7%) | ≥0.03922 |

| | | | | | | | | | | |
|---|---|---|---|---|---|---|---|---|---|---|
| India | 64.4% (60.0%, 68.6%) | ≥20.48 | 48.5% (44.0%, 53.0%) | ≥0.048 | 74.0% (70.0%, 77.8%) | ≥24.91 | 49.1% (44.6%, 53.6%) | ≥0.1073 | 44.3% (39.8%, 48.8%) | ≥0.01176 |
| Tanzania | 47.6% (40.3%, 55.0%) | ≥8.997 | 64.0% (56.7%, 70.9%) | ≥0.127 | 55.6% (48.2%, 62.8%) | ≥3.28 | 40.2% (33.2%, 47.6%) | ≥0.0854 | 39.7% (32.7%, 47.0%) | ≥0.00392 |
| Madagascar | 73.5% (66.5%, 79.7%) | ≥47.8 | 78.4% (71.7%, 84.1%) | ≥0.777 | 81.6% (75.3%, 86.9%) | ≥47.98 | 78.4% (71.7%, 84.1%) | ≥0.3751 | 71.4% (64.3%, 77.7%) | ≥0.50196 |
| **Group** | | | | | | | | | | |
| Male | 73.0% (70.7%, 75.3%) | ≥43.66 | 69.0% (66.6%, 71.3%) | ≥0.382 | 67.2% (64.8%, 69.6%) | ≥10.39 | 65.8% (63.3%, 68.2%) | ≥0.2746 | 63.9% (61.4%, 66.3%) | ≥0.11765 |
| Female | 63.8% (61.3%, 66.2%) | ≥14.65 | 60.2% (57.7%, 62.7%) | ≥0.083 | 68.8% (66.4%, 71.1%) | ≥4.05 | 62.0% (59.5%, 64.4%) | ≥0.1339 | 58.3% (55.8%, 60.8%) | ≥0.01961 |
| HIV-positive | 46.0% (41.0%, 51.0%) | ≥9.061 | 53.0% (48.0%, 58.0%) | ≥0.069 | 52.0% (47.0%, 57.0%) | ≥2.35 | 33.9% (29.3%, 38.8%) | ≥0.0897 | 42.2% (37.3%, 47.2%) | ≥0.01176 |
| HIV-negative | 79.5% (77.8%, 81.0%) | ≥42.98 | 74.6% (72.9%, 76.3%) | ≥0.398 | 73.1% (71.3%, 74.8%) | ≥11.16 | 73.0% (71.2%, 74.7%) | ≥0.2941 | 68.3% (66.5%, 70.2%) | ≥0.09804 |
| Diabetes | 80.8% (74.6%, 86.0%) | ≥42.98 | 82.3% (76.3%, 87.4%) | ≥0.592 | 78.8% (72.4%, 84.3%) | ≥15.84 | 78.3% (71.9%, 83.8%) | ≥0.3498 | 71.7% (64.9%, 77.9%) | ≥0.11765 |
| No diabetes | 73.3% (71.5%, 75.1%) | ≥35.11 | 67.2% (65.4%, 69.1%) | ≥0.203 | 69.0% (67.1%, 70.8%) | ≥6.45 | 64.8% (62.9%, 66.7%) | ≥0.1915 | 64.6% (62.7%, 66.5%) | ≥0.0549 |

| | | | | | | | | | | |
|---|---|---|---|---|---|---|---|---|---|---|
| History of TB | 58.2% (54.1%, 62.2%) | ≥47.89 | 52.9% (48.8%, 57.0%) | ≥0.594 | 54.1% (50.0%, 58.2%) | ≥24.38 | 52.0% (47.9%, 56.2%) | ≥0.3498 | 48.0% (43.8%, 52.1%) | ≥0.25098 |
| No history of TB | 80.3% (78.5%, 82.0%) | ≥33.35 | 78.3% (76.5%, 80.1%) | ≥0.214 | 79.5% (77.7%, 81.2%) | ≥6.75 | 72.6% (70.6%, 74.5%) | ≥0.1924 | 73.8% (71.8%, 75.7%) | ≥0.0549 |

1. TB risk scores ranged from 0-100 for CAD4TB and INSIGHT CXR, and 0-1 for qXR, DrAid and Genki

**Supplemental Figure 1**. **Forest plot of the sensitivity and specificity of qXR by country and subgroup using a universal threshold**. (A) The sensitivity and specificity by country, with 95% CIs; (B) The sensitivity and specificity by subgroup, with 95% CIs. The overall accuracy of the CAD algorithm is listed at the bottom with a vertical dashed red line in order to compare the overall estimate to the country and subgroup estimates.

**A**

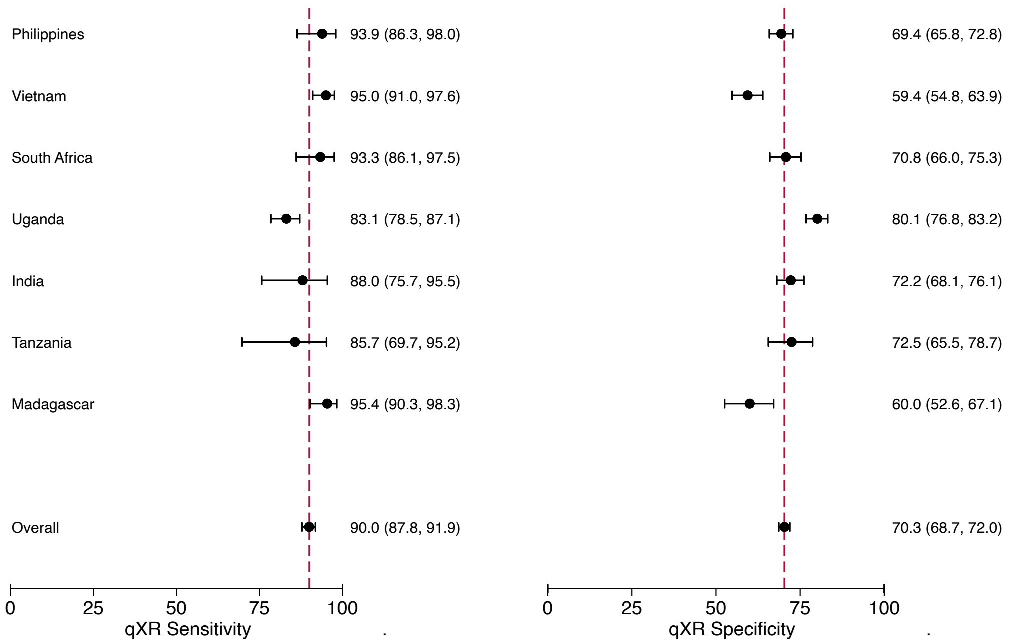


**B.**

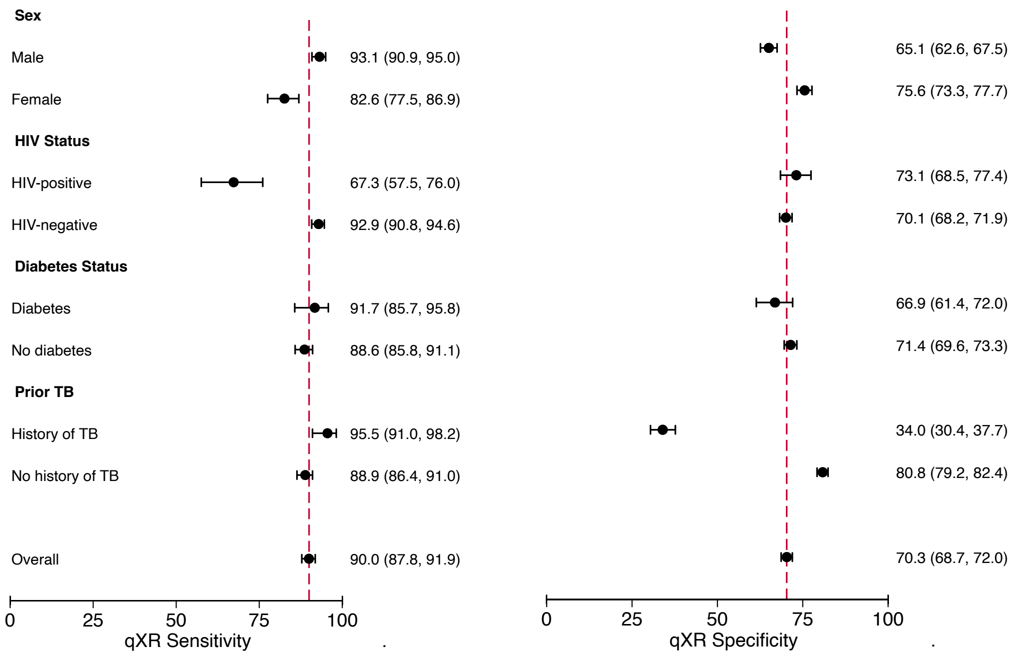

**Supplemental Figure 2**. **Forest plot of the sensitivity and specificity of INSIGHT CXR by country and subgroup using a universal threshold**. (A) The sensitivity and specificity by country, with 95% CIs; (B) The sensitivity and specificity by subgroup, with 95% CIs. The overall accuracy of the CAD algorithm is listed at the bottom with a vertical dashed red line in order to compare the overall estimate to the country and subgroup estimates.

**A**

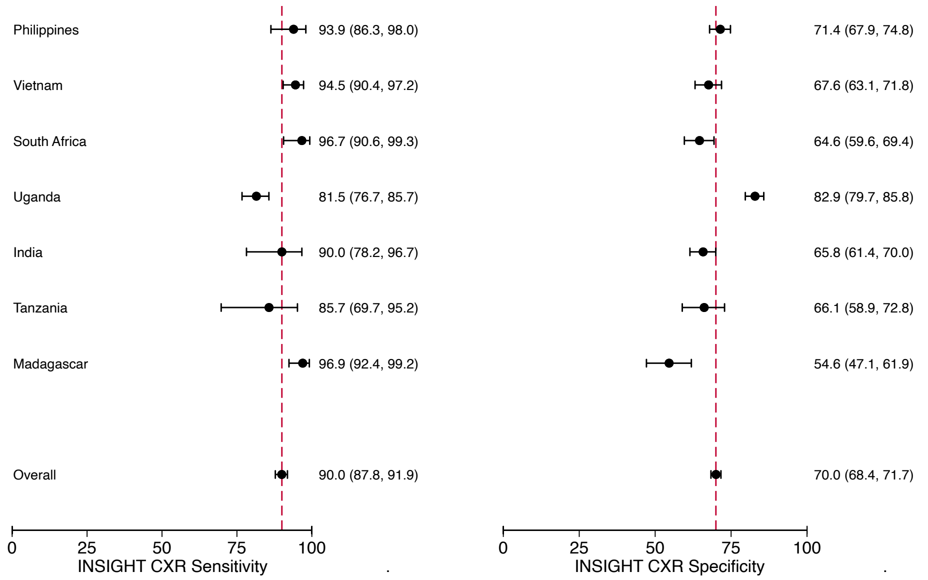


**B**

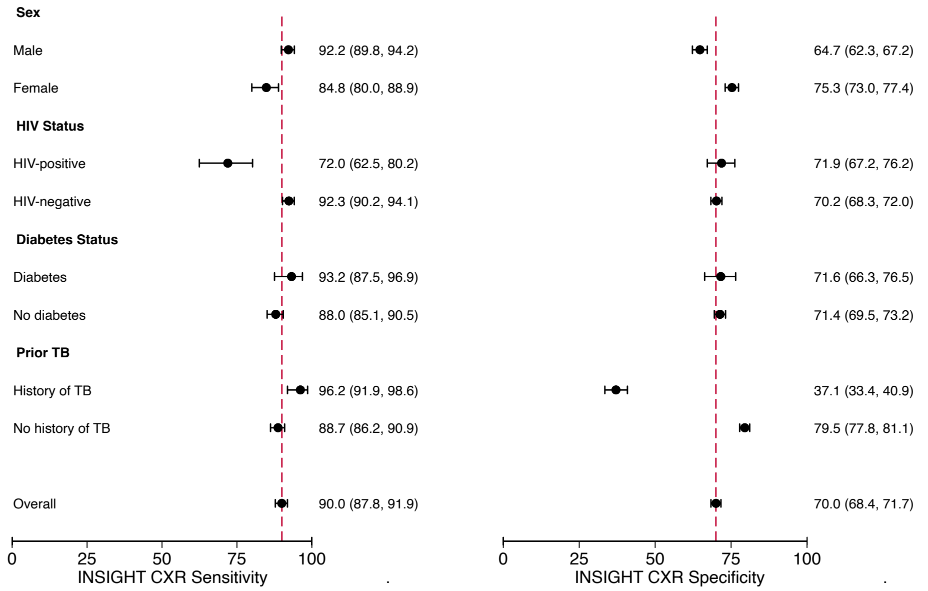

**Supplemental Figure 3**. **Forest plot of the sensitivity and specificity of DrAid by country and subgroup using a universal threshold**. (A) The sensitivity and specificity by country, with 95% CIs; (B) The sensitivity and specificity by subgroup, with 95% CIs. The overall accuracy of the CAD algorithm is listed at the bottom with a vertical dashed red line in order to compare the overall estimate to the country and subgroup estimates.

**A**

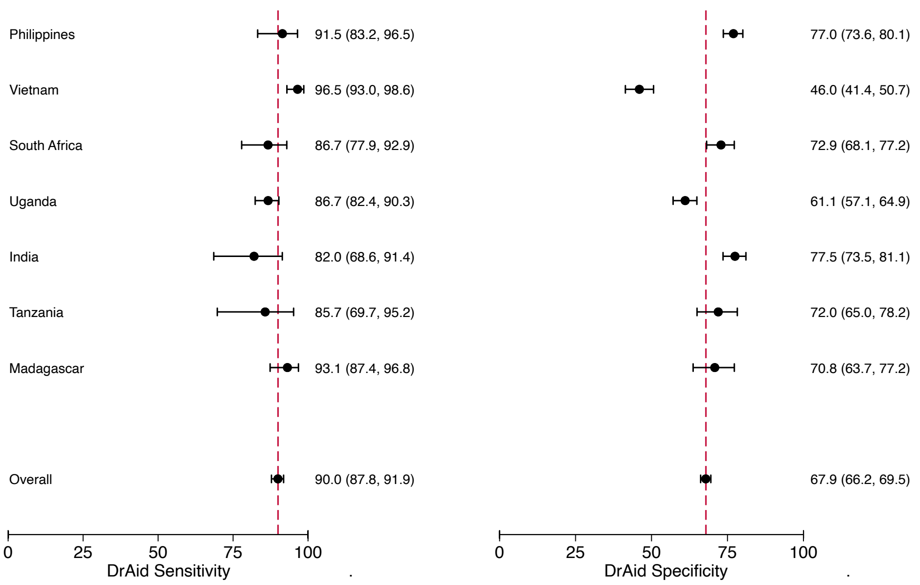


**B.**

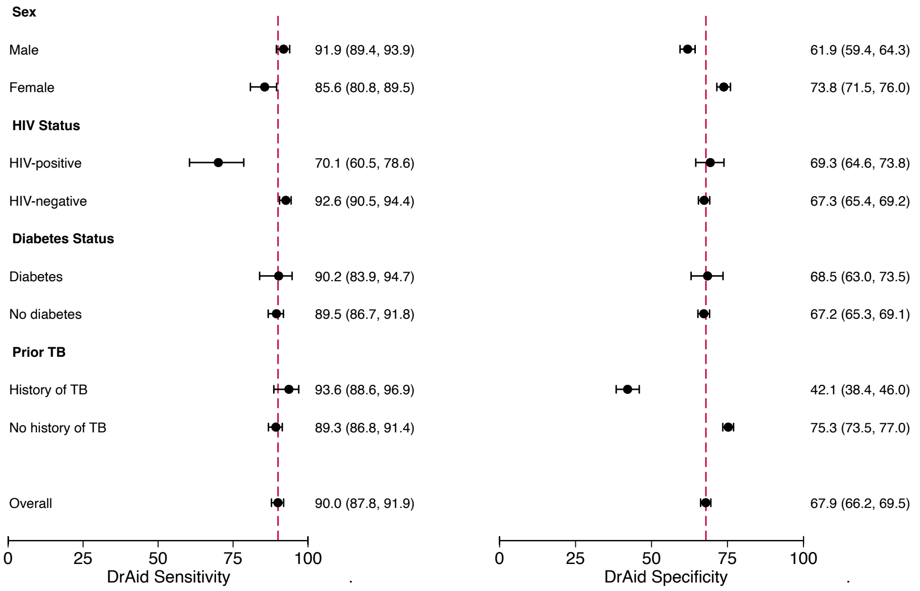

**Supplemental Figure 4**. **Forest plot of the sensitivity and specificity of Genki by country and subgroup using a universal threshold**. (A) The sensitivity and specificity by country, with 95% CIs; (B) The sensitivity and specificity by subgroup, with 95% CIs. The overall accuracy of the CAD algorithm is listed at the bottom with a vertical dashed red line in order to compare the overall estimate to the country and subgroup estimates.

**A.**

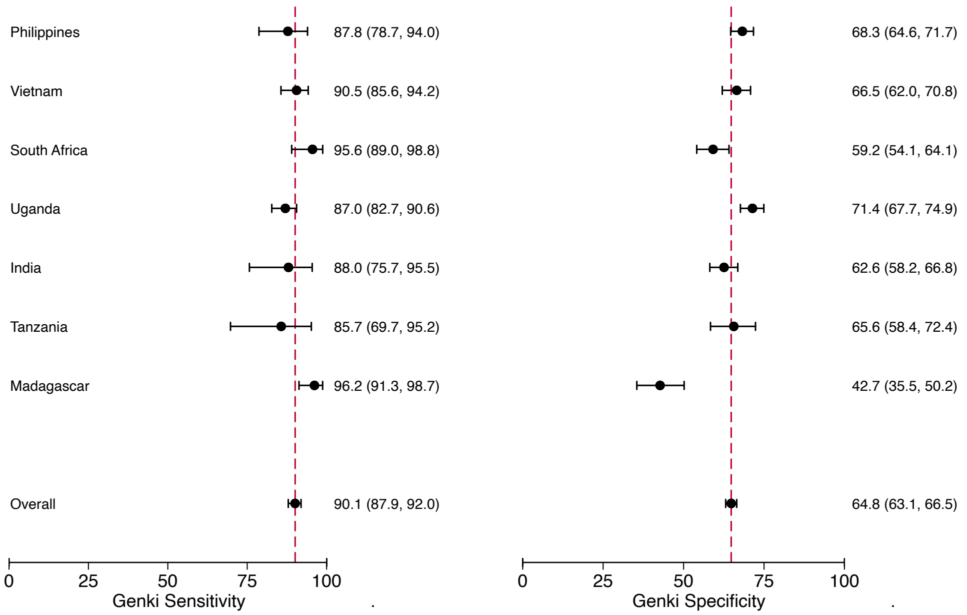


**B.**

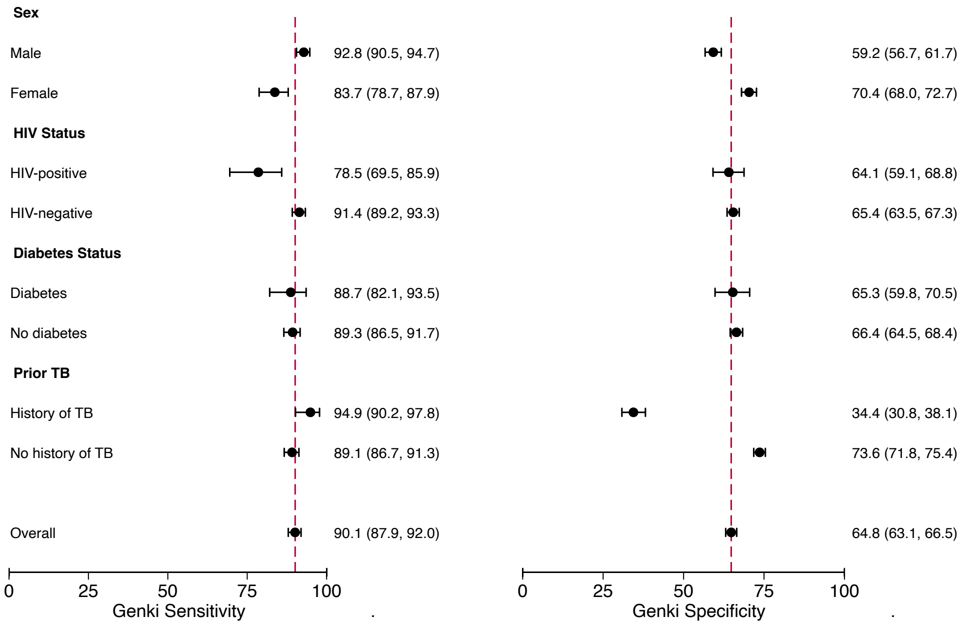
